# Linking international registries to FHIR and Phenopackets with RareLink: a scalable REDCap-based framework for rare disease data interoperability

**DOI:** 10.1101/2025.05.09.25327342

**Authors:** Adam S.L. Graefe, Filip Rehburg, Samer Alkarkoukly, Daniel Danis, Ana Grönke, Miriam R. Hübner, Alexander Bartschke, Thomas Debertshäuser, Sophie A.I. Klopfenstein, Julian Saß, Julia Fleck, Mirko Rehberg, Jana Zschüntzsch, Elisabeth F. Nyoungui, Tatiana Kalashnikova, Luis Murguía-Favela, Beata Derfalvi, Nicola A.M. Wright, Shahida Moosa, Soichi Ogishima, Oliver Semler, Susanna Wiegand, Peter Kuehnen, Christopher J. Mungall, Melissa A. Haendel, Peter N. Robinson, Sylvia Thun, Oya Beyan

## Abstract

While Research Electronic Data Capture (REDCap) has been widely adopted in rare disease research, its unconstrained data format often leads to implementations that lack native interoperability with global health data standards, limiting secondary data use. To address this, we developed and validated *RareLink*, an open-source framework implementing our previously-published ontology-based rare disease common data model, enabling standardised data exchange between REDCap, international registries, and downstream analysis tools. Its preconfigured pipelines interact with the local REDCap application programming interface and enable semi-automatic import or export of data to the Global Alliance for Genomics and Health (GA4GH) Phenopackets and Health Level 7 (HL7) Fast Healthcare Interoperability Resources (FHIR) instances, conforming to the HL7 International Patient Summary and Genomics Reporting profiles. The framework was developed in three iterative phases using retrospective and prospective clinical data from patients with various rare metabolic and neuromuscular disorders, as well as inborn errors of immunity. Phase one involved deployment across four German university hospitals for registry and data analysis purposes. Phase two integrated RareLink with the Canadian Inborn Errors of Immunity National Registry, enhancing extensibility. Phase three focuses on international implementation in South Africa and Japan to assess global scalability. Implementation feedback was continuously incorporated to validate outputs and improve usability. For evaluation purposes, we defined a simulated Kabuki syndrome cohort based on published cases and demonstrated data export to both Phenopackets and FHIR instances. RareLink can enhance the clinical utility of REDCap by enabling structured data analysis and interoperability. Its global applicability and open-source nature can support equitable rare disease research with the ultimate goal to improve patient care. Broader adoption and coordination with entities such as HL7 and the European Reference Networks are thus essential to realise its full potential. The framework and its documentation are freely available through <u>GitHub</u> and <u>Read the Docs</u>, respectively.

## Introduction

Rare diseases represent a significant global health challenge with over 260 million affected worldwide. In Europe, a condition is classified as rare when it affects fewer than 5 of 10,000 people^1^. More than 10,000 distinct rare diseases have been identified^2^, the majority of which are genetic in origin and first manifest during childhood^1^. Diagnostic delays are common, often spanning several years^3^, and most rare diseases currently lack causal treatments^4^. Despite their collective impact, the field continues to face substantial barriers, including limited data quality and availability, complicating diagnosis, optimal care, and research progress. Limited resources and heterogeneous, non-interoperable information systems compound these challenges, resulting in rare disease datasets not conforming to international data standards^5^.

Due to the inherent fragmentation of rare disease individuals across institutions and health systems, interoperability is essential for enabling consistent data interpretation and exchange^6^. Adoption of medical ontologies, Health Level 7 (HL7) Fast Healthcare Interoperability Resources (FHIR), and the Global Alliance for Genomics and Health (GA4GH) Phenopacket Schema designed by the Monarch Initiative enables reliable data exchange, supporting registries, secondary data use, and precision medicine^6–9^. However, many hospital information systems either lack rare disease-specific data or do not support the integration of these standards natively, posing barriers to clinical and translational use^10^. Research Electronic Data Capture (REDCap) is a globally adopted, web-based, no-cost software for non-profit organisations frequently used for data collection in research^11^ and rare disease registries^5^. Importantly, all data is stored on-site within the hosting institution’s servers, ensuring data sovereignty and compliance with applicable data protection regulations. While its flexibility in project and data schema definitions allows simple usage, it often results in unconstrained, project-specific data schemas that restrict interoperability, complicating alignment with international data standards, cross-registry data sharing, and federated downstream analyses^12^.

In our previous work, we developed an ontology-based rare disease common data model (RD-CDM) harmonising the European Rare Disease Registry Infrastructure Common Data Set, FHIR, and Phenopackets^13^. Building on this foundation, we now present RareLink, a novel REDCap-based framework that integrates this model by predefining data collection instruments with embedded ontology codes. RareLink is configured to enable automated export to FHIR resource instances and Phenopackets via a command-line-interface, supported by documentation that guides users through each step of the workflow. This study presents the iterative development and evaluation of RareLink along with the generated documentation and software across real-world settings in Germany, Canada, South Africa, and Japan, covering diverse use cases and a broad spectrum of rare diseases.

### Search strategy and selection criteria

REDCap is widely adopted in rare disease research across a range of use cases. To assess its role in supporting data standards for rare diseases, we searched PubMed from inception to May 2, 2025 for publications in English using the following two queries: 1) “REDCap” AND “rare diseases” AND (“FHIR” OR “Phenopackets”), and 2) “REDCap” AND “rare diseases” AND “interoperability“; both returned no results. A broader third search for “REDCap” AND “rare diseases” identified 24 studies (supplementary table 1), primarily involving disease-specific research, registries, and international consortia focused on specific rare disease groups. We additionally searched for “REDCap” AND (“FHIR” OR “Phenopackets”) to identify existing integrations. Of five studies (supplementary table 1) found, three linked REDCap with FHIR including the Clinical Data Interoperability Services module used by our framework. One introduced Convert-Pheno, a toolkit for translating between REDCap, Phenopackets and other models, though without a rare disease-specific and ontology-based model preconfigured within REDCap for FHIR and Phenopackets.

## Methods

### Overall project design

Our work on RareLink followed an iterative and collaborative development process over the past three years (figure 1). A diverse group of stakeholders contributed to its development and validation, providing domain-specific expertise at each stage. Clinical experts from metabolic, neuromuscular, immunodeficiency, clinical genetics, and other rare diseases guided the clinical applicability. Interoperability experts supported the implementation of ontologies and health data standards. Software developers and REDCap administrators contributed to the technical development of the framework, while doctoral and medical students assisted with data capture, analysis and iterative prototyping.

**Figure 1:**
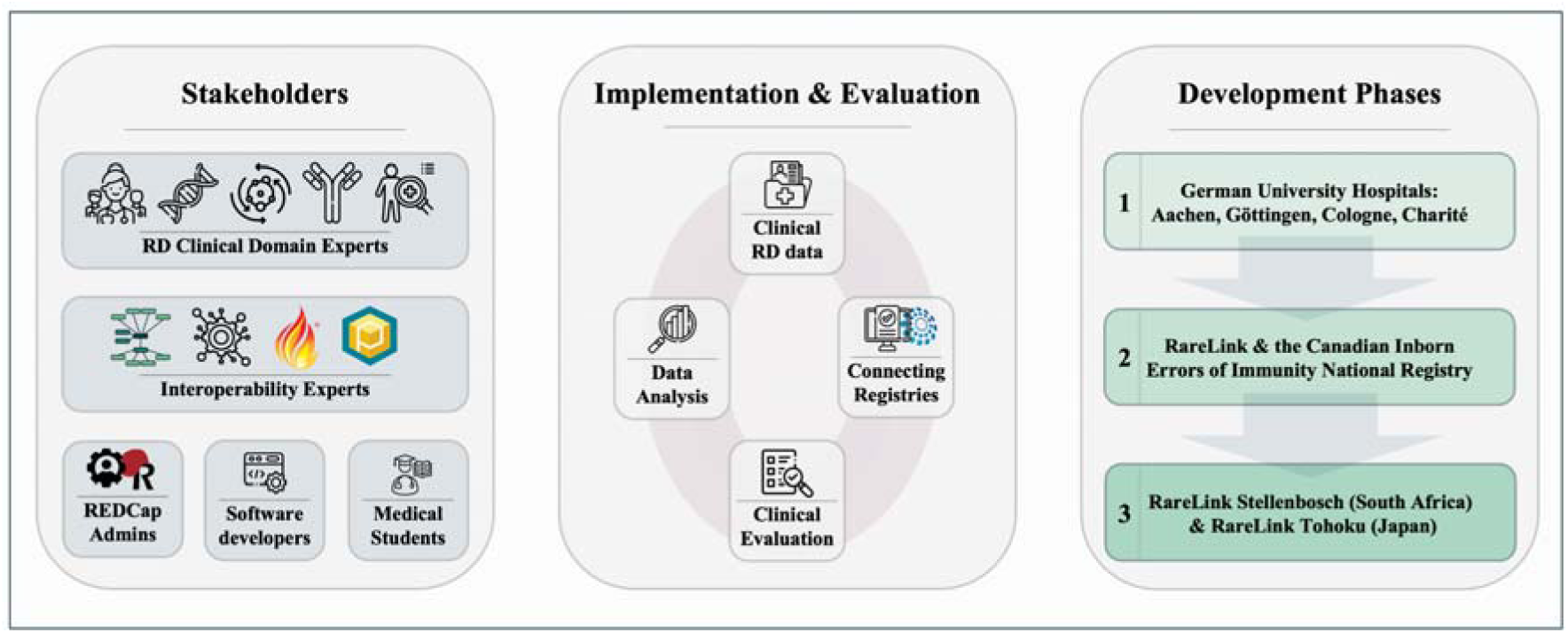
Development of the RareLink framework. This figure illustrates the methodological foundation of the RareLink framework, including key stakeholders, implementation, evaluation, and three development phases. Implementation involved clinical rare disease data from diverse sources further specified in the main text; applied for data analysis and registry linkage, including the European Reference Networks. Development proceeded through three overlapping phases: (1) deployment at German university hospitals in Aachen, Göttingen, Cologne, and Charité Berlin; (2) integration with the Canadian Inborn Errors of Immunity National Registry, and (3) implementation at Stellenbosch University (South Africa) and Tohoku University (Japan). RD=rare diseases.

### Technical development

Developed with Python 3.12 and an object-oriented design, the RareLink codebase is available on GitHub under an open-source Apache v2.0 license^14^. We have built several RareLink Python packages, including utilities, the Typer-based command line interface, and a RareLink-Phenopacket module. Additionally, a Sphinx-based RareLink documentation, linked to our GitHub repository, centralises all information required for the installation, usage, and implementation of the RareLink framework^14^.

We embedded the entire ontology-based RD-CDM^13^ into REDCap, with each section represented as one instrument. All ontologies associated with the data elements and value sets were incorporated in REDCap’s variable name and choices fields with their respective Internationalized Resource Identifiers prefixes while adhering with its naming constraints (i.e. lowercase letters, underscores only, and no additional special characters). Within REDCap’s Field Annotations, each element was defined along with its semantic codes and mappings to FHIR and Phenopackets, creating an integrated expression repository. By leveraging REDCap’s repeating instruments and required fields, we defined the cardinalities for our embedded data model to ensure compliance with the HL7 International Patient Summary (IPS) v2.0.0^15^, the HL7 Genomics Reporting v3.0.0^16^, and the GA4GH Phenopacket Schema v2.0^7^. The resulting REDCap data model and accompanying data dictionary, named *RareLink-CDM* (short for RareLink Common Data Model), was defined with LinkML^17^ v1.8.7, a unified data modeling language with integrated Python functionalities. We use this specification to process and validate exported REDCap records and leverage the LinkML-Map module to support semi-automated import of tabular data into REDCap. Detailed specifications of the RareLink-CDM are provided in the corresponding section of the RareLink documentation^14^.

Mapping and export of our RareLink-CDM to HL7 FHIR were performed using the open-source toFHIR engine^18^. We uploaded the FHIR v4.0.1 Base Resources, the Genomics Reporting v3.0.0, and IPS v2.0.0-ballot profiles into the aforementioned engine, ensuring that exports of the RareLink-CDM produce validated FHIR resource instances compliant with these resources and profiles. Additionally, we developed specific RareLink-CDM v2.0.0 profiles using the FHIR Shorthand (FSH) Language and the FSH compiler SUSHI Unshortens SHorthand Inputs (SUSHI) v3.15.0. A set of example instances were validated against the corresponding Structure Definitions and published using the IG Publisher v1.8.25.

The RareLink-Phenopacket module was designed to be compatible with other REDCap data models that are embedding ontologies. The Python package includes general mapping and creation functions for multiple Phenopacket blocks and Python classes for processing while integrating the Phenopackets Python package to ensure automatic validation. The mapping configuration for the RareLink-CDM was defined according to the underlying RD-CDM^13^. Further, we developed guidelines for defining other REDCap data models and their corresponding configurations that can also utilise the module^14^.

### Implementation and clinical applications

We deployed RareLink in multiple real-world settings to support both registry development and data analysis, with iterative refinements to enhance international scalability and semantic consistency. Implementations encompassed a range of rare diseases, including rare neuromuscular, neurometabolic, hereditary bone, and developmental disorders, inborn errors of immunity, and undiagnosed cases, using both retrospective and prospective clinical data extracted from the electronic health records (EHR) and local information systems.

Deployments were conducted in three overlapping phases, some of which are still ongoing. In phase one, RareLink was implemented at the four German university hospitals Berlin, Cologne, Cologne, Aachen, and Goettingen. Phenopacket-based analyses were performed, and data were captured for local registries and the European Reference Networks. Ethical approval was obtained from Berlin (approval number: EA2/242/22) and Cologne (approval number: 21-1477_1). In Cologne data was collected as part of the European Reference Network on rare bone diseases (ERN BOND). In Aachen data use was governed by institutional registry protocols under the responsibility of the respective clinical data custodians. In Göttingen data was captured as part of the EURO-NMD (Neuromuscular disorder) Registry^19^ (approval number: 3/2/25Ü). In phase two, we integrated RareLink with the Canadian Inborn Errors of Immunity National Registry (CIEINR)^20^, whose extended data model informed the design of additional domain-specific forms compatible with our framework with ethical approval numbers from the Izaak Walton Killam Hospital for Children in Halifax (IWK-REB:1028743) and the University of Calgary (UofC-REB:23-0108).

In phase three, RareLink is currently being implemented at Stellenbosch University (South Africa) and Tohoku University (Japan), with initial testing conducted using synthetic data and preparations underway for real-world clinical deployment within the Undiagnosed Disease Programme in South Africa^21^ (ethical approval number from Stellenbosch University Health Research Ethics Committee: N18/03/031) and partners in Japanese rare disease projects. While this study does not report detailed cohort-level findings, its focus was on evaluating global scalability, adapting to country-specific and international requirements, and refining the framework accordingly.

Feedback was continuously gathered throughout the iterative deployment process via informal exchanges, implementation meetings, and hands-on user testing. Rather than following a formalised evaluation protocol, feedback was integrated through a collaborative, back-and-forth process with clinical and technical partners. As such, the evaluation reflects a qualitative synthesis of observations and implementation experiences across diverse settings. To assess the semantic precision of the generated data, we created a simulated cohort of ten individuals with Kabuki Syndrome type 1 based on published cases and entered their data using our RareLink-CDM. In addition to genetic findings, data on the disease, phenotypic features, clinical measurements, and demographics were captured. We exported these records to both FHIR and Phenopacket formats and compared the output for semantic consistency. The complete evaluation cohort including its REDCap source data, Phenopackets, and FHIR instances are publicly available in our GitHub repository^14^.

## Results

The RareLink framework is an open-source, modular software designed for seamless integration with any REDCap instance connected to the BioPortal Ontology Services terminology server and a valid country-specific Systemized Nomenclature of Medicine – Clinical Terms (SNOMED CT) license (figure 2). RareLink consists of five core modules: (i) RareLink Documentation, (ii) RareLink Common Data Model, (iii) RareLink Command-Line Interface (CLI), (iv) RareLink FHIR Module, and (v) RareLink-Phenopackets Module. It is fully compatible with local REDCap projects and institutional deployments, supporting diverse use cases, including patient registries, study cohorts, and related clinical research activities. The framework is designed to be user-friendly for both clinical and technical personnel. By integrating the complete ontology-based RD-CDM^13^ within REDCap, RareLink enables guided manual data entry for prospective cohorts and semi-automatic import using LinkML-Map^17^ for retrospective data sets. Once data is captured, users can export to validated HL7 FHIR instances and GA4GH Phenopackets by using the CLI that enables them to interact with the toFHIR Module and the RareLink-Phenopackets Module (figure 2). The Clinical Data Interoperability Services (CDIS) module^22^ can be integrated to import existing FHIR instances to REDCap.

**Figure 2:**
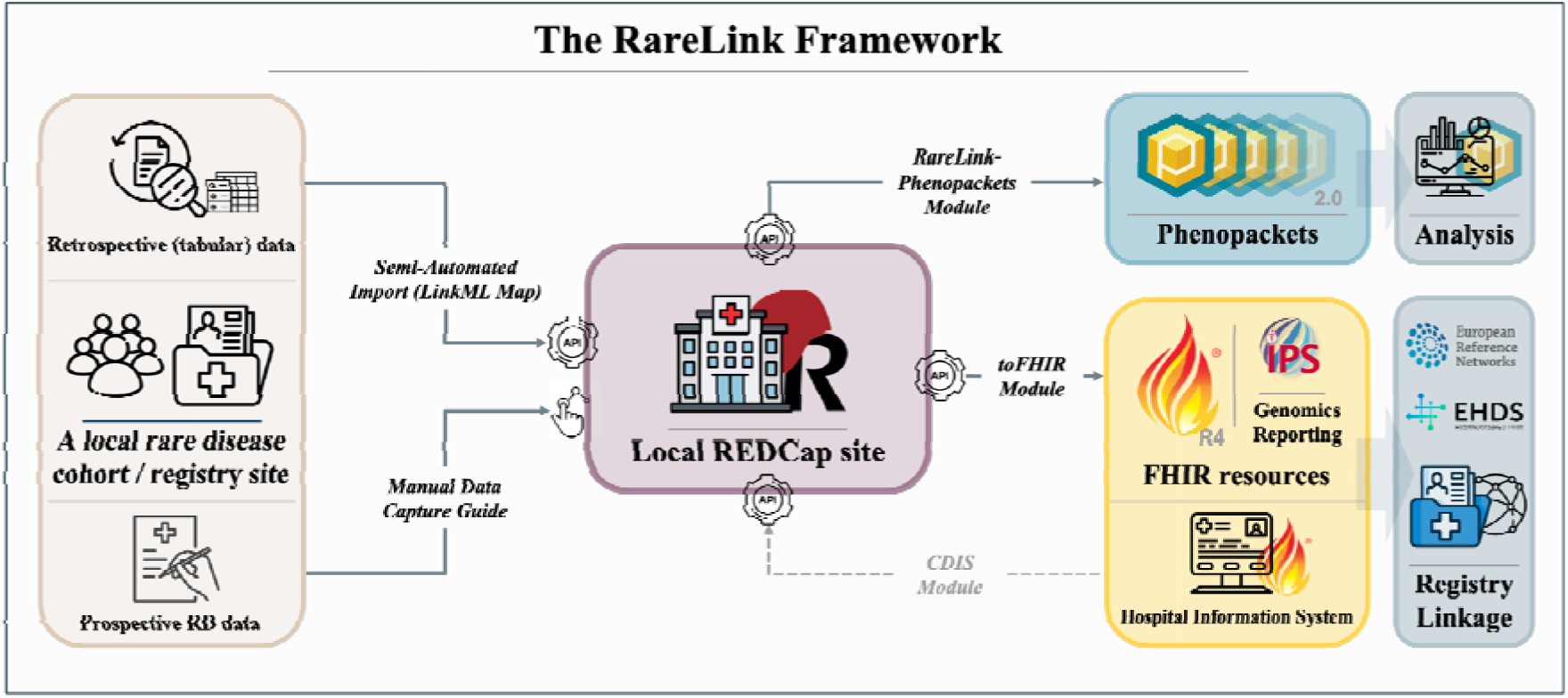
Overview of the entire RareLink framework’s data flow. The RareLink framework is integrated with a local REDCap instance and preconfigured for the RareLink common data model. Utilising the local REDCap API and the RareLink-CLI, the toFHIR & CDIS modules can export to HL7 FHIR International Patient Summary (IPS) and Genomic Reporting resources and import from a corresponding FHIR server enabling export or record linkage to the European Reference Networks, the EHDS, or other international and domain-specific registries. The RareLink-Phenopackets module exports to GA4GH Phenopackets, allowing for the use of Phenopacket-based analysis software. Additionally, the LinkML-based import mapper supports data import from tabular databases into REDCap, and the Manual Data Capture Guide aids with the manual entry of data according to the common data model. API=Application Programming Interface. CDIS=Clinical Data Interoperability Services. CLI=Command Line Interface. EHDS=European Health Data Space. FHIR=Fast Healthcare Interoperability Resources. GA4GH=Global Alliance for Genomics and Health. IPS=International Patient Summary. RareLink documentation=https://rarelink.readthedocs.io/en/latest/index.html.

### RareLink Documentation

The comprehensive RareLink documentation was designed with an emphasis on reusability, scalability, and cross-institutional applicability. Given RareLink’s deployment across multiple countries and institutions, clear and accessible documentation was essential to ensure consistent implementation and independent use. It centralises all RareLink features, guides, and resources, accessible from any REDCap site, and is structured into five subsections: Background, RareLink Framework, Installation, User Guide, and Additional Information. The *Background* section provides theoretical context and introduces all employed ontologies with references for further reading. The *RareLink Framework* section offers an overview of the architecture, including the RareLink-CDM and the command-line interface. *Installation* guides users through framework setup, REDCap configuration, data dictionary import, and REDCap API connection. Detailed user guides support manual data capture, semi-automated data entry, use of the Phenopackets and FHIR modules, development of REDCap instruments, and REDCap tools integration. The *Additional Information* section includes contribution guidelines, a changelog, FAQs, acknowledgements, licensing, and contact information (https://rarelink.readthedocs.io/en/latest/index.html).

### RareLink common data model

We enhanced the ontology-based RD-CDM^13^ for its REDCap implementation by developing the RareLink Common Data Model (RareLink-CDM), aligning it with corresponding FHIR resources and profiles, and Phenopacket blocks. To ensure interoperability, we defined required fields within each section and extended the Measurements section to support linkage with International Patient Summary (IPS) profiles for laboratory, imaging, and procedural data. Only the Formal Criteria section is mandatory; all other sections are optional but conditionally required: if any field is used, its associated mandatory elements must be completed. With exception of the Formal Criteria, Personal Information, Patient Status, Consent, and Disability, all sections are implemented as repeating instruments (figure 3A-C).

**Figure 3:**
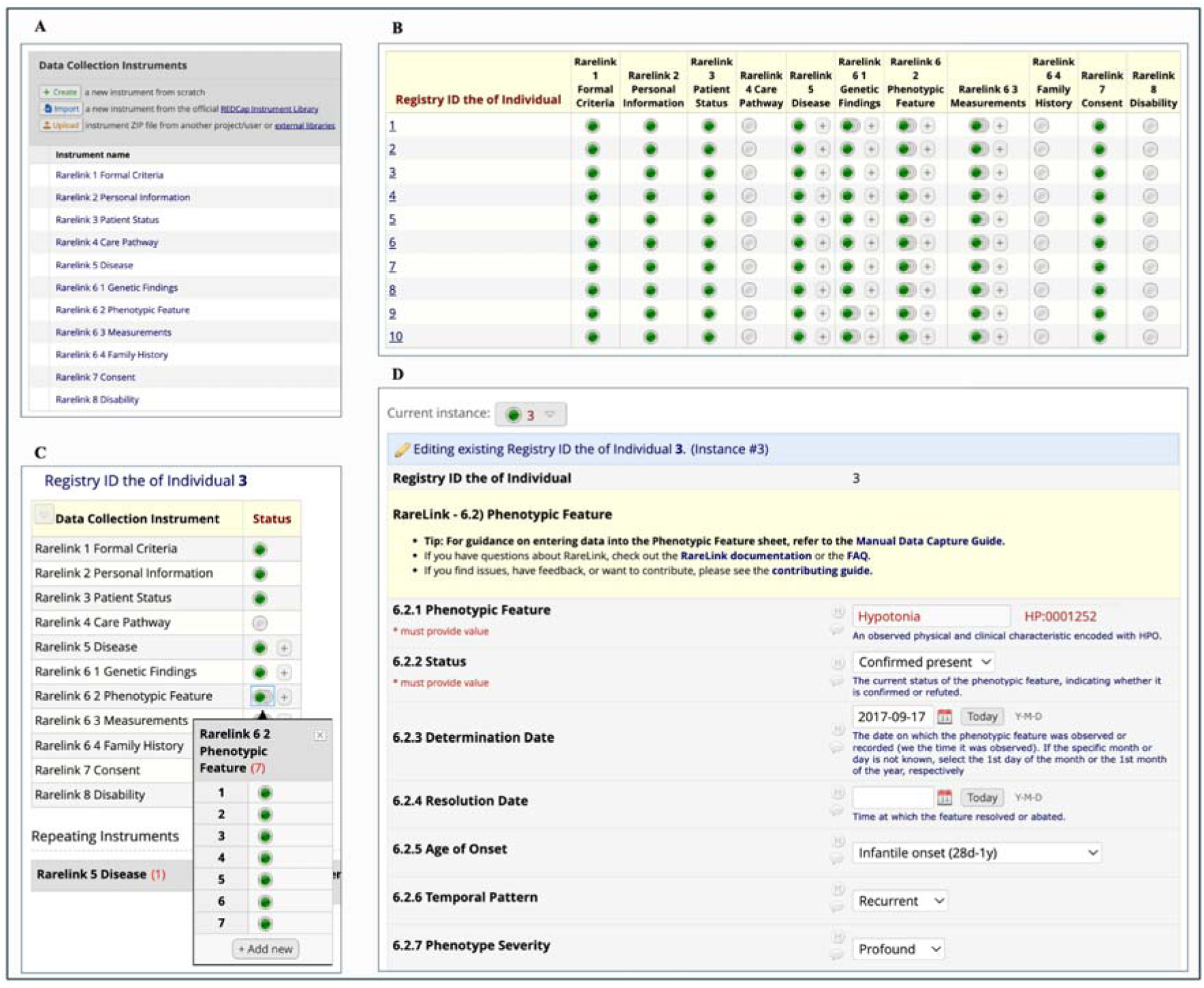
The RareLink-CDM in REDCap for the evaluation cohort of ten individuals with Kabuki syndrome type 1. A) Overview of all RareLink-CDM sections displayed as standalone data collection instruments within REDCap’s Designer view. B) The record status dashboard displaying the ten simulated individuals enrolled in the registry, each with completed data across demographic, consent, disease, genetic, phenotypic, and measurement instruments. C) The record dashboard for individual 3, illustrating the use of repeating instances for phenotypic features. D) A screenshot of the data entry window for a phenotypic feature of individual 3, including introductory text and links to the documentation and manual data capture guide. In this example, the individual was reported with confirmed, recurrent, and profound hypotonia, with an infantile onset dated 17 September 2017. HP=Human Phenotype Ontology.

To promote accurate data capture and reduce entry errors, the REDCap instruments include detailed instructions, branching logic, and embedded links, complemented by the manual data capture guide (figure 3D). The LinkML representation of the RareLink-CDM mirrors this structure by grouping repeating instances in its JSON serialisation. Once data is exported as REDCap-JSON, it is automatically processed and validated against the LinkML-JSON schema. This schema^14^, along with the corresponding Python classes, includes all REDCap variables, coded terms, and value sets as importable enumerations.

### Command Line Interface

The RareLink command-line interface (CLI) guides users through the entire workflow from setup to data export, with a design that accommodates clinicians with limited coding experience. It features a user-friendly interface with descriptive commands, embedded links, and contextual hints. Each command includes cross-references and built-in validation to ensure required steps are completed and configurations are correct. Organised into five command groups, the CLI facilitates onboarding and streamline usage (figure 4A). After setup, the *redcap* command group enables API-based interaction with local REDCap projects. The *fhir* and *phenopackets* command groups provide all necessary steps for exporting data in both formats (figure 4B). As adoption of RareLink grows, additional commands will be incorporated based on community-driven feedback to support evolving needs.

**Figure 4:**
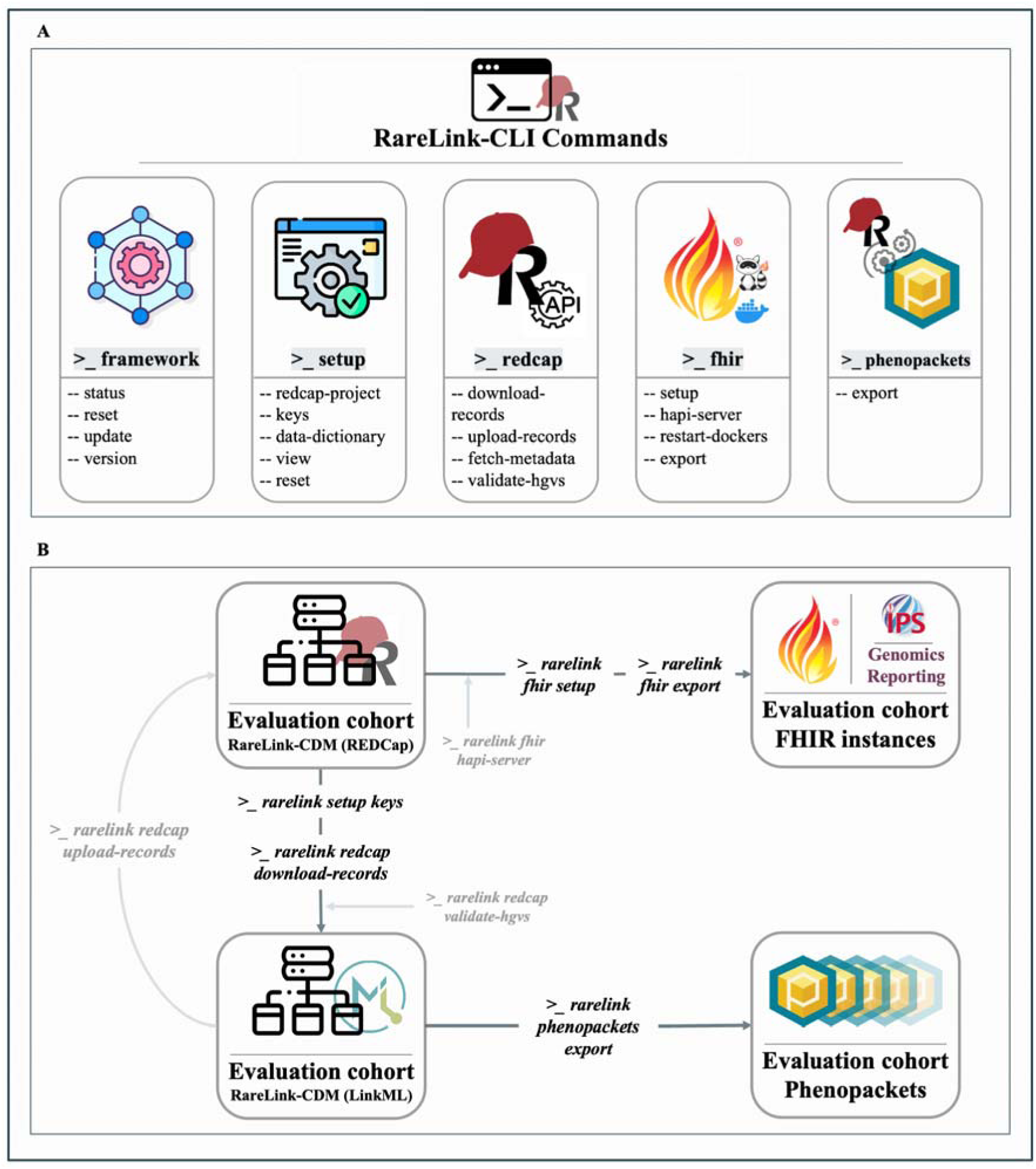
RareLink command-line interface and data flow for the Kabuki Type 1 evaluation cohort. A) The RareLink CLI is organised into five primary command groups - *framework* (global configuration), *setup* (local installation), *redcap* (interaction with a REDCap project), *fhir* (FHIR export via the RareLink-FHIR module) and *phenopackets* (Phenopacket generation via the RareLink-Phenopackets module) - with further subcommands under each. Future releases will extend functionality as requirements evolve. B) Key data-flow steps (*download-records*, *phenopackets export*, *fhir export*) are invoked after setup (*setup keys*, *fhir setup,* and, if necessary, *fhir hapi-server* and *redcap validate-hgvs)* on the CLI to fetch, validate, and transform REDCap data into the LinkML representation of the RareLink-CDM, Phenopackets, and FHIR instances (see supplementary figure 1 for full console output). The LinkML data can be imported back into REDCap using the *redcap upload-records* command. CDM=common data model. FHIR=Fast Healthcare Interoperability Resources. LinkML=unified data modeling language. REDCap=Research Electronic Data Capture.

### RareLink-FHIR-Module

All 75 elements mapped to FHIR in the ontology-based RD-CDM^13^ are exportable from the RareLink-CDM to the corresponding FHIR resources and profiles that we selected. The export process leverages the toFHIR engine, which enables automatic validation against the IPS v2.0.0 profiles, the Genomics Reporting v3.0.0 profiles, and Base Resources v4.0.1 structure definitions to any FHIR server (figure 5). These profiles are embedded as dependencies within the RareLink-CDM profiles ensuring interoperability upon implementation. This functionality facilitates linkage to FHIR repositories, international registries, and supports data import via the Clinical Data Interoperability Services module^22^. The export pipeline requires Docker to be installed and running. In the absence of a remote FHIR server, a local instance can be set up using a HAPI server. Detailed guidance and all CLI commands are provided in the user guide documentation under the FHIR module section^14^. The RareLink-CDM FHIR specifications are publicly available and hosted through our repository^14^.

**Figure 5.**
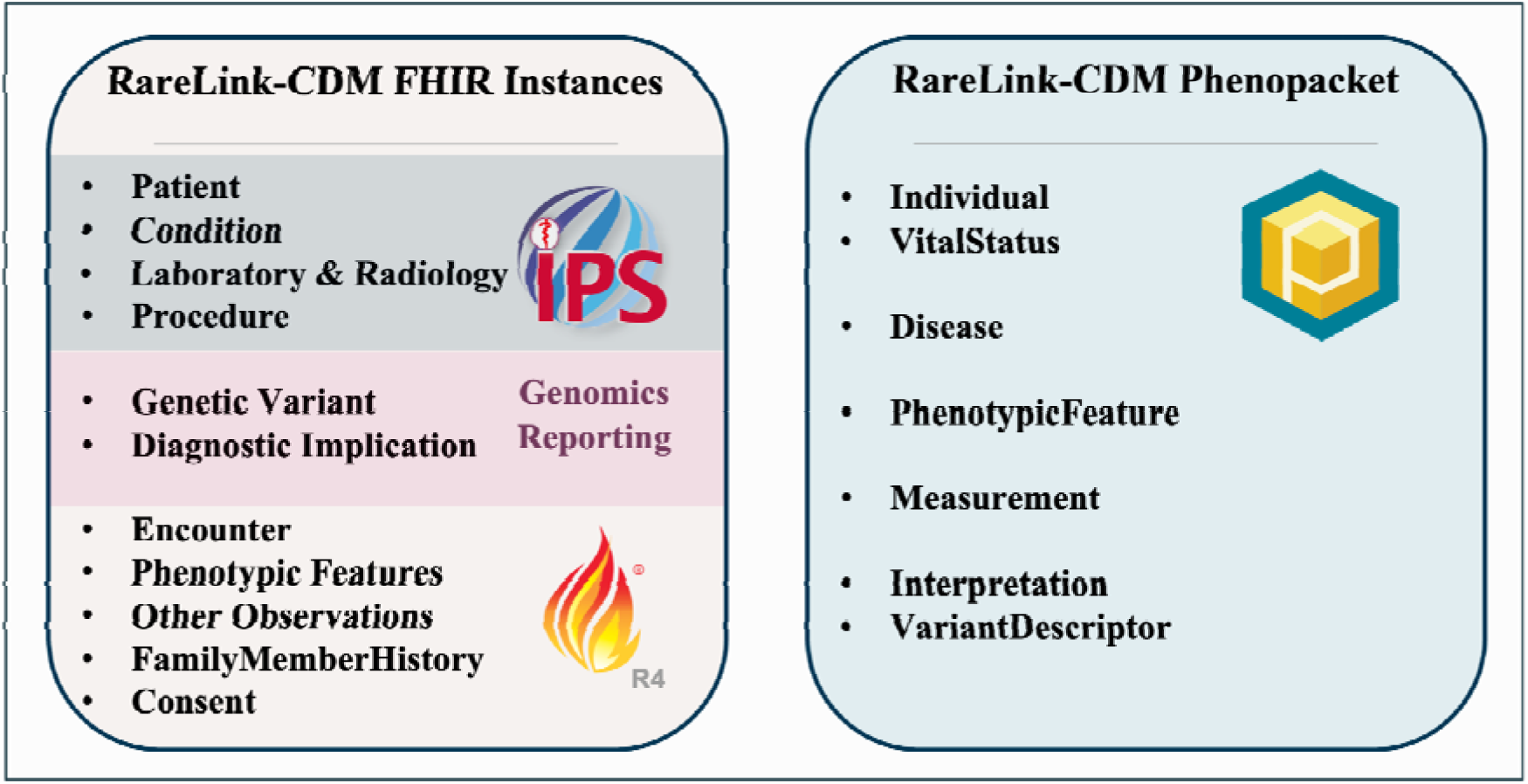
Schematic overview of the RareLink-CDM as both FHIR instances and a Phenopacket. FHIR instances conform to the HL7 International Patient Summary v2.0.0 profiles for Patient, Condition, Laboratory, Radiology, and Procedure. Genetic findings are captured using the HL7 Genomics Reporting v3.0.0 Genetic Variant and Diagnostic Implication profiles. Additional components, including encounters, phenotypic, and other observations (e.g. age category, gestational age), family history and consent (incorporating ERDRI-CDS elements), utilise FHIR R4 base resources. The RareLink-CDM Phenopacket comprises Individual, VitalStatus, and Disease blocks, together with phenotypic and measurement data, and genetic information within the Interpretation and VariantDescriptor blocks. ERDRI-CDS=European Rare Disease Infrastructure Common Data Set. HL7=Health Level 7. IPS=International Patient Summary. RareLink-CDM=RareLink Common Data Model.

### RareLink-Phenopackets-Module

With the exception of the family history section, all 43 elements mapped to Phenopackets in the ontology-based RD-CDM^13^ are exportable from REDCap via the RareLink-CDM in the current version (figure 5). Following export, the dataset is processed and validated against the LinkML schema, after which the CLI *phenopackets export* command generates the corresponding Phenopackets (figure 4C). A free-of-charge Bioportal API key is required to retrieve ontology labels during this process. The export engine leverages RareLink’s DataProcessor class to convert REDCap codes into valid *OntologyClass* elements within the Phenopacket structure. Mapping logic is defined in the BaseMapper class to support the necessary Phenopacket blocks. By integrating mapping, creation, writing, and validation functionalities within a single pipeline, the engine streamlines the export process. While the mappings are preconfigured for the RareLink-CDM, they remain extensible. Developers seeking to adapt the Phenopacket engine for other data models should follow RareLink’s guidelines for building ontology-based REDCap instruments. Detailed setup instructions for extending the engine are provided in the Phenopackets Module section of the documentation^14^.

## Clinical Applications

### Clinical usability

Evaluation of the RareLink framework demonstrated usability and interoperability across diverse REDCap environments. During testing, users highlighted the intuitive documentation, the user-friendly CLI, and automated export pipelines as key strengths. The system successfully supported the export and analysis of all elements defined in the ontology-based RD-CDM that were mapped to either FHIR or Phenopackets, with the sole exception of the family history section in Phenopackets^13^, demonstrating its adaptability and coverage. Notably, predefining cohort-specific elements improved usability and consistency, facilitating subsequent registry deployment and data analysis. Its integration into the adopted REDCap infrastructure was noted as a major advantage, offering a scalable solution as demonstrated in phase two and three. However, REDCap’s typical separation from hospital information systems was identified as a barrier to seamless integration with clinical workflows. While the framework’s modular design and extensive guidance support its reusability across various research settings, the evaluation also highlighted challenges for users without a background in coding or interoperability. Clinicians unfamiliar with command-line tools required additional support to operate the framework effectively. Further, the local semantic annotation requires substantial understanding of ontologies and terminologies. To support these issues, feedback mechanisms embedded in the documentation and GitHub repository^14^ are actively collecting input for future enhancements.

### Canadian Inborn Errors of Immunity National Registry

The RareLink framework demonstrated both scalability and extensibility through its application in the CIEINR^20^. The collaboration with CIEINR served as a key use case for evaluating extensibility and scalability. In this domain-specific implementation, the core RareLink-CDM was extended with additional clinical fields tailored to immunodeficiency-specific needs. Drawing from this experience, we developed guidelines for creating REDCap instruments compatible with the RareLink framework and provided detailed documentation on adapting the modular Phenopacket module for export. Iterative trial data capture, continuous feedback loops, and refinement processes informed these developments. Evaluation showed that the core RareLink-CDM sufficiently covered key data elements, such as demographics, patient clinical status, and genetic information. Additional domain-specific sections, such as phenotypic features, were incorporated through modular extensions and controlled value sets. Mandatory elements were preserved to maintain interoperability, and rule-based validation ensured consistency across the extended model. The CIEINR is in the implementation stage and has been proven operational for Phenopackets in the piloting phase. Specific development and analysis on the use of the RareLink framework will be reported elsewhere. This use case confirmed that RareLink can be adapted for diverse clinical domains while preserving semantic and syntactic integrity.

### Evaluation cohort

The evaluation cohort comprised ten simulated individuals with Kabuki Syndrome type 1. For each case, data included basic demographics, phenotypic features, genetic findings and selected clinical measurements. Following data entry, records were exported to both FHIR instances and Phenopackets (figure 4B). The resulting JSON files are publicly available via our GitHub repository^14^. As illustrated in supplementary figure 2, the exported data maintain consistent semantic representations across formats for each individual, while ensuring syntactic interoperability with the respective data standard

## Discussion

In this study, we present RareLink, an open-source framework built around REDCap that enhances interoperability for rare disease research, studies, and registries. Our framework integrates the ontology-based RD-CDM^13^ into REDCap, facilitating its usage across countries and use-cases. RareLink provides predefined REDCap instruments and extension guides that enable direct export to Phenopackets and FHIR resource instances. In addition, it supports both guided manual data capture and, via LinkML, semi-automated import of retrospective cohorts. To support usability and global update, we developed a centralised, openly accessible documentation online and a command-line interface for seamless interaction with local REDCap instances.

RareLink can notably enhance data precision and availability for large cohorts and international registries, while also supporting downstream and advanced analyses. Recent studies employing large cohorts have significantly advanced our understanding of rare diseases while highlighting the need for precise encoding and interoperability^23–25^. The European Reference Networks^26^ and the European Health Data Space^27^ hold promise for harmonising extensive rare disease data. Yet, many disease-specific registries contain highly-specialised data incompatible with FHIR or Phenopackets. RareLink can seamlessly connect the European Rare Disease Infrastructure Registry Common Data Set^28^ to these data standards through a ready-to-use implementation. Its FHIR profiles inherit those of the HL7 IPS and Genomics Reporting and could therefore be beneficial for global rare disease efforts^29^. Moreover, the integration with Phenopackets facilitates the use of reusable analysis pipelines, such as the novel GPSEA (genotype-phenotype: statistical evaluation of associations)^30^ or PhEval (Phenotypic inference Evaluation framework)^31^.

Extending the RareLink-CDM model with the CIEINR^20^ provided valuable insights into establishing domain-specific common data elements^32^ that are compatible with our framework. The use of ontologies has been essential for preserving medical semantics while ensuring compatibility with the given data standards and other domain-specific organisations^8^. Although RareLink has proven effective for multi-site research due to its lightweight implementation around any REDCap instance, its success will ultimately rely on organisational interoperability and governance within each use case.

Important limitations to highlight include inherent constraints of REDCap. It is typically not integrated within clinical workflows or the EHR^22^, which can necessitate additional resources and limit the effectiveness of a common data model^33^. Although a local RareLink installation can import FHIR data more easily via the Clinical Data Interoperability Service module^22^ using the RareLink-CDM profiles, linking existing patient records remains challenging, with difficulties varying by the underlying information system^22^. Moreover, license-related issues may arise with REDCap, the requirement for BioPortal connectivity, and the need for a country-specific SNOMED CT license.

Another limitation relates to the novelty of the data model^13^ implemented into RareLink. While it incorporates the European Rare Disease Registry Infrastructure Common Data Set^28^ and links to HL7 FHIR and the GA4GH Phenopacket Schema, to our knowledge, this study represents its first implementation with real world data.. Consequently, even though a direct connection with FHIR and Phenopackets is ensured, the introduction of another common data model may contribute to further fragmentation among cohorts^33^. Moreover, the RareLink-CDM FHIR profiles and Implementation Guide remain in draft trial-use status and subject to formal governance—including versioning, conformance testing, stakeholder review, and the incorporation of changes arising from the ballot process—before achieving formal approval. Finally, country-specific or multi-language implementation challenges highlight the indispensable need for coordinated efforts with international stakeholders, such as HL7, the IPS, the European Reference Networks, and the GA4GH^33^.

Future research should also leverage advanced FHIR infrastructures and features, such as SMART-on-FHIR^34^ and FHIR Questionnaires^35^ to enhance integration with EHRs and apply RareLink’s specifications to other clinical information systems. This would position Phenopackets closer to routine clinical data, thereby increasing available data for precision medicine and federated analyses. Moreover, broader implementation of RareLink across additional use cases, cohorts, and countries is warranted. Ultimately, extensive feedback will be essential to refine the documentation, engine, command-line interface, and specifications, thus improving adaptability and overall utility.

In conclusion, RareLink enhances data precision for rare disease experts by leveraging the widely adopted REDCap system. Its preconfigured pipelines and international accessibility make it a valuable resource for researchers. By enabling standardised analysis algorithms^30,31^ and linking modern registries^26^, RareLink can significantly advance our understanding and treatment of rare diseases. Its effectiveness, however, centers on the community-driven and open-source effort. Only through such collaborative engagement, RareLink can evolve into a reliable global resource for rare disease research in REDCap, supporting equity regardless of financial or geographic constraints.

## Contributors

A.S.L.G. conceived the study and led the manuscript writing and development of all framework modules under the supervision of P.N.R., O.B., and S.T. In phase one, J.F., M.R., O.S., S.W., P.K., J.Z., and E.N. managed local data capture for their respective registries and assisted in data analysis. Phase two in Canada was supported by T.K., L.M., B.D., and N.A.M.W. Phase three activities in South Africa and Japan were carried out with S.M. and S.O., respectively. F.R., D.D., and P.N.R. oversaw the development of all Phenopackets-related components, while S.A., A.G., M.H., A.B., T.D., S.A.I.K., J.S., and S.T. contributed to the design and implementation of FHIR–related modules. C.J.M. and M.A.H. integrated LinkML and ontology frameworks into RareLink. Finally, C.J.M., M.A.H., P.N.R., S.T., and O.B. provided guidance on the overall conceptual design. All authors reviewed and approved the final manuscript.

## Data Sharing

Supplementary material appendix [link].

The full RareLink framework is available via the RareLink GitHub repository (https://github.com/BIH-CEI/RareLink). Comprehensive documentation is accessible at https://rarelink.readthedocs.io/en/latest/index.html. The fictional evaluation cohort data are likewise available within the repository (https://github.com/BIH-CEI/rarelink/tree/develop/res/evaluation_cohort). The RareLink implementation guide and the corresponding RareLink-CDM profiles are available under https://bih-cei.github.io/rarelink/.

## Acknowledgments

We thank Jael Cheng for data capture in Cologne and Mostafa Kamal for assistance with the local REDCap instance. In Aachen, we are grateful to Zahra Mirkhasli and Thomas Eggermann for their support with local data capture. We also acknowledge Anil Sinaci and Doğukan Çavdaroğlu (SRDC toFHIR) for their assistance with development and implementation of their engine. We appreciate the contributions of Bernt Popp, Kai Heitmann, Annic Weyersberg, and Shahim Essaid in providing clinical context and addressing interoperability challenges, Niclas Worthmann for his work on graphical design, and Sinan Akalin for assistance on software development. Finally, we thank Yolandi Swart (Stellenbosch University), Brenda Turley (University of Calgary), and Taylor Mattinson (Dalhousie University) for their support during the respective site implementations.

