## Supplementary Material for "Linking international registries to FHIR and Phenopackets with RareLink: a scalable REDCap-based framework for rare disease data interoperability"

| **Table 1. Studies identified in the search strategy with titles highlighted in bold (May 2nd 2025)** | |
|---|---|
| **PubMed Search** | **Citation** |
| | Rashid R, Copelli S, Silverstein JC, Becich MJ. R**EDCap and the National Mesothelioma Virtual Bank—a scalable and sustainable model for rare disease biorepositories**. Journal of the American Medical Informatics Association. 2023 Oct 1;30(10):1634-44. |
| | Della Casa F, Vitale A, Lopalco G, Ruscitti P, Ciccia F, Emmi G, Cattalini M, Wiesik-Szewczyk E, Maggio MC, Ogunjimi B, Sfikakis PP. **Development and implementation of the AIDA international registry for patients with undifferentiated systemic autoinflammatory diseases**. Frontiers in medicine. 2022 Jun 10;9:908501. |
| | Nagy A, Molay F, Hargadon S, Brito Pires C, Grant N, De La Rosa Abreu L, Chen JY, D'Souza P, Macnamara E, Tifft C, Becker C. **The spectrum of neurological presentation in individuals affected by TBL1XR1 gene defects.** Orphanet Journal of Rare Diseases. 2024 Feb 20;19(1):79. |
| | Gaggiano C, Vitale A, Tufan A, Ragab G, Aragona E, Wiesik-Szewczyk E, Ait-Idir D, Conti G, Iezzi L, Maggio MC, Cattalini M. **The autoinflammatory diseases alliance registry of monogenic autoinflammatory diseases. Frontiers in Medicine**. 2022 Sep 9;9:980679. |
| | Durkin N, Pellegrini M, Gorter R, Slater G, Cross KM, Ure B, Wijnen R, Gottrand F, Eaton S, De Coppi P, ERNICA. **Follow-up and transition practices in esophageal atresia: a review of European Reference Network on rare Inherited and Congenital Anomalies (ERNICA) centres and affiliates**. Pediatric Surgery International. 2024 Nov 9;40(1):300. |
| | Vitale A, Caggiano V, Della Casa F, Hernández-Rodríguez J, Frassi M, Monti S, Tufan A, Telesca S, Conticini E, Ragab G, Lopalco G. **Development and implementation of the AIDA international registry for patients with VEXAS syndrome**. Frontiers in Medicine. 2022 Jul 11;9:926500. |
| | Alzarka B, Morizono H, Bollman JW, Kim D, Guay-Woodford LM. **Design and implementation of the hepatorenal fibrocystic disease core center clinical database: a centralized resource for characterizing autosomal recessive polycystic kidney disease and other hepatorenal fibrocystic diseases**. Frontiers in pediatrics. 2017 Apr 20;5:80. |
| "REDCap" AND "rare diseases" | Vitale A, Della Casa F, Ragab G, Almaghlouth IA, Lopalco G, Pereira RM, Guerriero S, Govoni M, Sfikakis PP, Giacomelli R, Ciccia F. **Development and implementation of the AIDA International Registry for patients with Behçet's disease**. Internal and emergency medicine. 2022 Oct;17(7):1977-86. |
| | Casa FD, Vitale A, PEREIRA RM, Guerriero S, Ragab G, Lopalco G, Cattalini M, Mattioli I, Parronchi P, Paroli MP, GIUDICE ED. **Development and Implementation of the AIDA International Registry for Patients with Non-Infectious Scleritis.** Ophthalmology and Therapy. 2022. |
| | Della Casa F, Vitale A, Guerriero S, Sota J, Cimaz R, Ragab G, Ruscitti P, Pereira RM, Minoia F, Del Giudice E, Emmi G. **Development and implementation of the AIDA international registry for patients with non-infectious uveitis.** Ophthalmology and Therapy. 2022 Jan 31;11(2):899. |
| | Vitale A, Della Casa F, Lopalco G, Pereira RM, Ruscitti P, Giacomelli R, Ragab G, La Torre F, Bartoloni E, Del Giudice E, Lomater C. **Development and implementation of the AIDA international registry for patients with Still's disease.** Frontiers in Medicine. 2022 Apr 7;9:878797. |
| | Seed L, Scott A, Pichini A, Peter M, Tadros S, da Costa CS, Hill M. **Perceptions of genomic newborn screening: a cross-sectional survey conducted with UK medical students.** BMJ open. 2024 Sep 1;14(9):e089108. |
| | Nunes FL, Ferriani MP, Moreno AS, Langer SS, Maia LS, Ferraro MF, Sarti W, Bessa Junior JD, Cunha D, Suffritti C, Dias MM. **Decreasing attacks and improving quality of life through a systematic management program for patients with hereditary angioedema.** International Archives of Allergy and Immunology. 2021 Aug 2;182(8):697-708. |
| | Szövérfi Z, Lazáry Á, Varga PP. **Primary Spinal Tumor Registry in the National Centre for Spinal Disorders.** Orvosi Hetilap. 2014 May 1;155(19):745-9. |
| | Sota J, Vitale A, Więsik-Szewczyk E, Frassi M, Lopalco G, Emmi G, Govoni M, de Paulis A, Marino A, Gidaro A, Monti S. **Development and implementation of the AIDA international registry for patients with Schnitzler's syndrome.** Frontiers in medicine. 2022 Jul 18;9:931189. |
| | Matsuki K, Harada-Shiba M, Hori M, Ogura M, Akiyama Y, Fujii H, Ishibashi Y, Ishida T, Ishigaki Y, Kabata D, |

| | |
|---|---|
| “REDCap” AND “rare diseases” | Kihara Y. **Association between familial hypercholesterolemia and serum levels of cholesterol synthesis and absorption markers: the CACHE study FH analysis**. Journal of atherosclerosis and thrombosis. 2023 Sep 1;30(9):1152-64. |
| | Della Casa F, Vitale A, Cattalini M, La Torre F, Capozio G, Del Giudice E, Maggio MC, Conti G, Alessio M, Ogunjimi B, Ragab G. **Development and implementation of the AIDA International Registry for patients with Periodic Fever, Aphthous stomatitis, Pharyngitis, and cervical Adenitis syndrome.** Frontiers in pediatrics. 2022 Jul 22;10:930305. |
| | Glassberg JA, Linton EA, Burson K, Hendershot T, Telfair J, Kanter J, Gordeuk VR, King AA, Melvin CL, Shah N, Hankins JS. **Publication of data collection forms from NHLBI funded sickle cell disease implementation consortium (SCDIC) registry.** Orphanet journal of rare diseases. 2020 Dec;15:1-6. |
| | MacMullen LE, George-Sankoh I, Stanley K, McCormick EM, Muraresku CC, Goldstein A, Zolkipli-Cunningham Z, Falk MJ. **Bridging the clinical-research gap: harnessing an electronic data capture, integration, and visualization platform to systematically assess prospective patient-reported outcomes in mitochondrial medicine**. Molecular Genetics and Metabolism. 2024 May 1;142(1):108348. |
| | Hiremath G, Kodroff E, Strobel MJ, Scott M, Book W, Reidy C, Kyle S, Mack D, Sable K, Abonia P, Spergel J. **Individuals affected by eosinophilic gastrointestinal disorders have complex unmet needs and frequently experience unique barriers to care**. Clinics and research in hepatology and gastroenterology. 2018 Oct 1;42(5):483-93. |
| | Perry DC, Arch B, Appelbe D, Francis P, Spowart C, Knight M. The BOSS Study. **Determining the incidence and clinical outcomes of uncommon conditions and events in orthopaedic surgery.** Bone & Joint Open. 2020 Mar 18;1(3):41-6. |
| | Li JQ, Dell J, Höller T, Fink D, Schmid M, Heinz C, Finger RP. **The treatment exit options for uveitis (TOFU) registry: Involving patients in the generation of evidence.** Gesundheitswesen (Bundesverband der Arzte des Offentlichen Gesundheitsdienstes (Germany)). 2021 Nov 3;83(S 01):S39-44. |
| | Mansoorshahi S, Scurlock C, Research Registry SA, Prakash SK. **Methodological advances in patient-centered rare disease research: the UTHealth Houston Turner Syndrome Society of the United States research registry.** Orphanet Journal of Rare Diseases. 2024 Mar 11;19(1):112. |
| | Velasco Puyo P, Christou S, Campisi S, Rodríguez-Sánchez MA, Reidel S, Perez-Hoyo S, Mota M, Savvidou I, Rekleiti A, Salvo A, Voi V. **COVID-19 in patients affected by red blood cell disorders, results from the European registry ERN-EuroBloodNet**. Orphanet Journal of Rare Diseases. 2025 Apr 16;20(1):183. |
| "REDCap" AND ("FHIR" OR "Phenopackets") | Cheng AC, Duda SN, Taylor R, Delacqua F, Lewis AA, Bosler T, Johnson KB, Harris PA. **REDCap on FHIR: clinical data interoperability services. Journal of biomedical informatics.** 2021 Sep 1;121:103871. |
| | Metke-Jimenez A, Hansen D. FHIRCap: **Transforming REDCap forms into FHIR resources.** AMIA Summits on Translational Science Proceedings. 2019 May 6;2019:54. |
| | Rueda M, Leist IC, Gut IG. **Convert-pheno: a software toolkit for the interconversion of standard data models for phenotypic data**. Journal of Biomedical Informatics. 2024 Jan 1;149:104558. |
| | Stäubert S, Strübing A, Schmidt F, Yahiaoui-Doktor M, Reusche M, Meineke F, Neumann D, Loeffler M. **The Concept of a Versatile Computing Tool Chain for Utilizing the Core Data Set of the Medical Informatics Initiative in the INTERPOLAR Project.** InGerman Medical Data Sciences 2024 2024 (pp. 59-66). IOS Press. |
| | Furner B, Cheng A, Desai AV, Benedetti DJ, Friedman DL, Wyatt KD, Watkins M, Volchenboum SL, Cohn SL. **Extracting electronic health record neuroblastoma treatment data with high fidelity using the REDCap clinical data interoperability services module.** JCO Clinical Cancer Informatics. 2024 May;8:e2400009. |

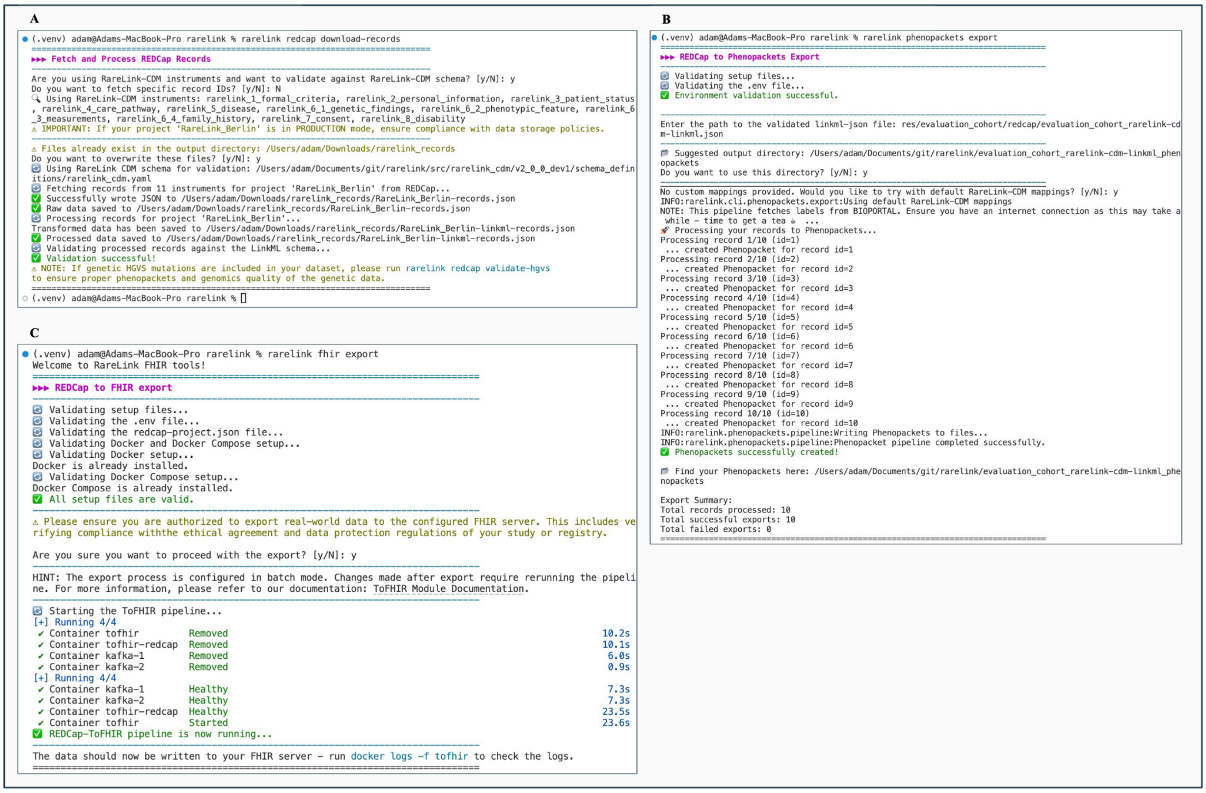


***Supplementary Figure 1:* Complete console output illustrating the data-processing workflow for the evaluation cohort.** A) The *rarelink redcap download-records* command retrieves all records from the local REDCap instance and validates them against the RareLink-CDM LinkML schema. B) The *rarelink phenopackets export* command transforms validated LinkML data into GA4GH Phenopackets, resolving ontology labels via BioPortal. C) The *rarelink fhir export* command invokes the toFHIR engine in Docker to convert REDCap data directly into FHIR R4 resources. CDM=common data model. CLI=command-line interface. FHIR=Fast Healthcare Interoperability Resources. LinkML=Linked Data Modeling Language.

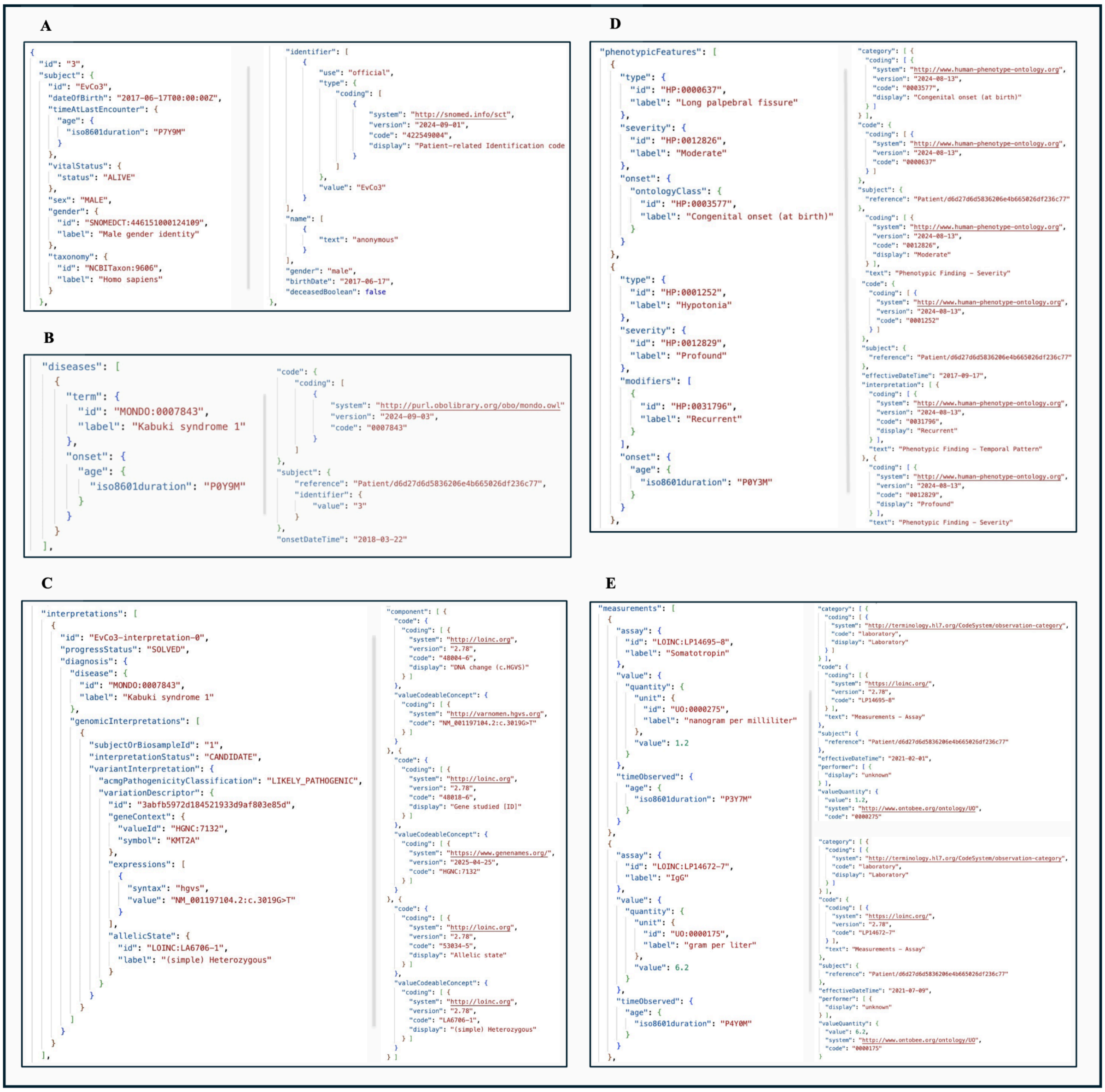


***Supplementary Figure 2*****: Representative JSON excerpts from a Phenopacket (left part in each subfigure) and corresponding FHIR resources (right part in each subfigure) for a single individual enrolled in the Kabuki Syndrome 1 evaluation cohort illustrating patient demographics, disease diagnosis, genetic findings, phenotypic features, and clinical laboratory measurements.** (A) The individual is a male (REDCap record ID: 3; cohort-specific ID: EvCo3), born on 2017-06-17 and at the last encounter alive. agnosis of Kabuki Syndrome 1 was made with an onset date of 2018-03-22, corresponding to an age of 9 months. (C) Genetic analysis identified a likely pathogenic, heterozygous variant in the KMT2A gene associated with the diagnosis. (D) Phenotypic presentation included a moderately expressed long palpebral fissure with congenital onset and recurrent profound hypotonia manifesting at 3 months of age. (E) A somatotropin concentration of 1.2 ng/mL was recorded on 2021-02-01, when the individual was 3 years and 7 months old, and an IgG concentration of 6.2 g/l was recorded on 2021-07-09, when the individual was 11 years and 4 months old. The complete set of synthetic test data used in this evaluation cohort is available in our public GitHub repository (https://github.com/BIH-CEI/rarelink/tree/develop/res/evaluation_cohort).
